# Epidemiological Development of Novel Coronavirus Pneumonia in China and Its Forecast

**DOI:** 10.1101/2020.02.21.20026229

**Authors:** Shanshan Wu, Panpan Sun, Ruiling Li, Liang Zhao, Yanli Wang, Lifang Jiang, Haili Wang, Jinbo Deng

## Abstract

**BACKGROUND AND OBJECTIVE:** The novel coronavirus (SARS-Cov-2) infected coronavirus disease 2019 (COVID-19) was broken out in Wuhan and Hubei province for more than a month. It severely threats people’s health of thousands in Chin and even other countries. In order to prevent its wide spread, it is necessary to understand the development of the epidemic with precise mathematical language.

**METHODS:** The various data of novel coronavirus pneumonia were collected from the official websites of the National Health Committee of the People’s Republic of China. According to epidemic and administrative division, three groups were divided to analyze the data, Hubei Province (including Wuhan), nationwide without Hubei and Henan Province. With classic SIR models, the fitting epidemiological curves of incidence have made, and basic reproduction number (R_0_) was also calculated as well. Therefore the disease’s infection intensity, peak time and the epidemiological end time can be deduced.

**RESULTS:** (1) Wuhan was the origin place of the epidemic, then it spread to Hubei province quickly. The patients in Hubei had increased rapidly with exponential rise. According to data in Hubei province, the fitting parabolas were made, and some with 51,673 cases. R_0_ curve shows with S-curve, at early breakout, R_0_ was as high as 6.27, then it decrease gradually. It is expected to approach to zero in early May; (2) In the group of nationwide without Hubei, the patient cases were much lower than Hubei, but its epidemiological fitting curve also shows a parabola as Hubei. The peak will arrive around February 10 with 9,145 cases. At beginning, R_0_ was as high as 2.44, then it decreases gradually and approach to zero in the end of March. (3) In Henan Province, the incidence stays very low, the parabolic fitting curve is similar to the nationwide without Hubei. The epidemic is expected to reach the peak on around February 12 and end in early April.

**CONCLUSION:** The epidemic development in all three groups shows parabolic curves. Their incidences are expected to reach their peaks on February 18 in Hubei, on February 10 in other areas of China. The epidemic will end in early May in Hubei, and in early April in other areas of China. Our study may provide useful knowledge for the government to make prevention and treatment policies.

## Introduction

The novel coronavirus pneumonia caused by SARS-CoV-2 is a highly infectious epidemic. The patients show systemic immunity decrease severely and strong respiratory symptoms with high death rate.^1^ The epidemic was first discovered in Wuhan, Hubei Province with strong infectiousness through multiple transmission routes, and some cases even show without any typical symptoms, bringing us a big problem for its prevention and control.^2 3^ World Health Organization (WHO) officially listed it as a “public health emergency of international concern”^4^ By February 16, 2020, China had confirmed 70548 patients cumulatively. Its so quick spread benefited from homecoming during Spring Festival, about 5 million people ever left Wuhan, the origin of COVID-19’s outbreak, for hometown before the city was closed on January 23, and 9 million people still remained in Wuhan. In these way, the 5 million people became source of infection to other ares in China. On the other hand, the disease broke out among the 9 million people in closed Wuhan. The epidemic has severely affected people’s lives and national economy. Over the past month, medical workers, scientists and governments adopted a serial powerful measures to treat patients and stop the epidemic spread. However, the number of infections is still growing in Wuhan and other areas of China, and there is a tendency to expand to the other countries.^5 6^ In order to prevent its widespread outward, it is necessary for us to understand the epidemiological development of the epidemic with a mathematical model. Recently, MedRxiv has published patients’ R0 value, it estimated R0=3.77 (95% CI: 3.51-4.05) in suspected patients and 3.00 (95%CI:2.82-3.20)in confirmed patients.7 In our study, using classic Susceptible Infected Recovered (SIR) soft, according current epidemic data, some important parameters, such as infection coefficient, removal coefficient and basic reproduction coefficient, were calculated. Than the fitting curves of incidence were drawn with peak time and end time of the epidemic. In addition, the dynamic values of R_0_ (Basic Reproduction Number) were also described as well. The several curves can help us to forecast the epidemic development. It will be useful for us to take targeted interventions to control the epidemic as soon as possible.

## Methods

All data came from the official websites of National Centers for Disease Control and National Health Committee of the People’s Republic of China. The confirmed cases were diagnosed according to the criteria of the COVID-19’s diagnoses and treatments, which were suggested by National Health Committee,^8 9^ including history of exposure, clinical symptoms (especially CT imaging) and positive nucleic acid test. Data were grouped into three according to their administrative division and epidemic condition as well: (1) Hubei Province (including Wuhan city), the group was defined, since it was the outbreak place of the disease and the closed province to prevent epidemic spread; (2) China without Hubei or nationwide except Hubei, it was set up, since its epidemic was infected by 5 million people from Wuhan before the closure of Wuhan city; (3) Henan Province, it was defined since Henan Province is the most populous province in China, with more than 100 million population, and frequent population exchanges with Wuhan.

Basic reproduction number (R_0_) was calculated.^10^ R_0_ refers to how many persons can be infected by a patient. If R_0_> 1, the disease will spread away soon and radially; R_0_ <1 means the infection is in controllable situation. So, 1 is regarded as an important threshold to distinguish whether the disease will spread or not.^11^ During the epidemiological development of the disease, R_0_ is changeable. The dynamic reproduction number is called R_t_ (Effective Reproduction Number). In order to prevent and control epidemic, we must find a way to reduce R_t_ below 1 as much as possible. Chain growth rate was also calculated in various groups. Generally it refers to a growth rate relative to previous period. The formula is: Chain growth rate = (value in current period-value in last period) / (value of last period)x100%. Chain growth rate represents the growth speed of the data. The larger chain growth rate is, the greater growth rate is. The negative value represents a negative growth rate.

Excel soft was used to record data and calculate some parameters, such as incidence and chain growth rate. In the meantime, using MATLAB software and SIR mathematical model, the parabolic epidemiological curves were simulated according to data. In fact, the fitting curve is tried to imitate the reality as much as possible. In addition, the curves of R_t_ were drawn as well. Finally, results were compared among three groups, Hubei Province, China without Hubei Province and Henan Province. P values were calculated as well, and P<0.05 refers to statistical signification.

## Results

According to original data, actual scatter plots were made. In fact, in early COVID-19 outbreak, either patient number or incidence increased with exponential curve in all three groups (table 1). For instance, from January 20 to February 17, the cases increased quickly from 270 to 59989 in Hubei province, and equivalently the incidence rise from 0.46⨯10^−5^ to 101.38⨯10^−5^. In Henan province, the cases increased from 0.00 to 1257, equivalently the incidences rise from 0.00 to 1.31⨯10^−5^. In the meantime, in nationwide without Hubei (China without Hubei), the incidences rise from 0 to 0.93⨯10^−5^. Compared with Hubei, in nationwide without Hubei has been maintained at low level, but epidemiological curve showed exponential increase similar to Hubei province. The cumulative incidences rate and chain growth rates are showed in figure 1 and figure 2. The incidence rate increased in Hubei province, compared with other area, and the increased speed in Hubei was faster than other areas in China.

**Table 1.** Cases, incidence and chain growth rate on various dates

| Date | Hubei |  |  | China without Hubei |  |  | Henan |  |  |
| --- | --- | --- | --- | --- | --- | --- | --- | --- | --- |
| | cases | Incidence<br>($1/10^5$) | Chain<br>growth<br>rate (%) | cases | Incidence<br>($1/10^5$) | Chain<br>growth<br>rate (%) | cases | Incidence<br>($1/10^5$) | Chain<br>growth<br>rate (%) |
| 1/20 | 270 | 0.46 | - | 21 | 0.00 | - | 0 | 0.00 | - |
| 1/22 | 444 | 0.75 | 18.40 | 127 | 0.01 | 95.38 | 5 | 0.01 | 400.00 |
| 1/24 | 729 | 1.23 | 32.79 | 558 | 0.04 | 98.58 | 32 | 0.03 | 255.56 |
| 1/26 | 1423 | 2.40 | 35.27 | 1321 | 0.10 | 43.12 | 128 | 0.13 | 54.22 |
| 1/28 | 3554 | 6.01 | 30.95 | 2420 | 0.18 | 34.37 | 206 | 0.21 | 22.62 |
| 1/30 | 5806 | 9.81 | 26.60 | 3886 | 0.29 | 24.35 | 352 | 0.37 | 26.62 |
| 2/1 | 9074 | 15.34 | 26.86 | 5306 | 0.40 | 14.40 | 493 | 0.51 | 16.82 |
| 2/3 | 13522 | 22.85 | 20.98 | 6916 | 0.52 | 14.73 | 675 | 0.70 | 19.26 |
| 2/5 | 19665 | 33.23 | 17.91 | 8353 | 0.63 | 9.25 | 851 | 0.89 | 11.39 |
| 2/7 | 24953 | 42.17 | 12.85 | 9593 | 0.72 | 6.01 | 981 | 1.02 | 7.33 |
| 2/9 | 29631 | 50.08 | 9.34 | 10540 | 0.79 | 4.38 | 1073 | 1.12 | 3.87 |
| 2/11 | 33366 | 56.39 | 5.16 | 11287 | 0.84 | 3.46 | 1135 | 1.18 | 2.71 |
| 2/13 | 51986 | 87.86 | 7.84 | 11865 | 0.89 | 2.30 | 1184 | 1.23 | 1.28 |
| 2/15 | 56249 | 95.06 | 3.39 | 12251 | 0.92 | 1.37 | 1231 | 1.28 | 1.57 |
| 2/17 | 59989 | 101.38 | 3.11 | 12447 | 0.93 | 0.66 | 1257 | 1.31 | 0.88 |
| 2/19 | 62031 | 104.84 | 0.57 | 12545 | 0.94 | 0.34 | 1265 | 1.32 | 0.24 |

**Fig 1.**
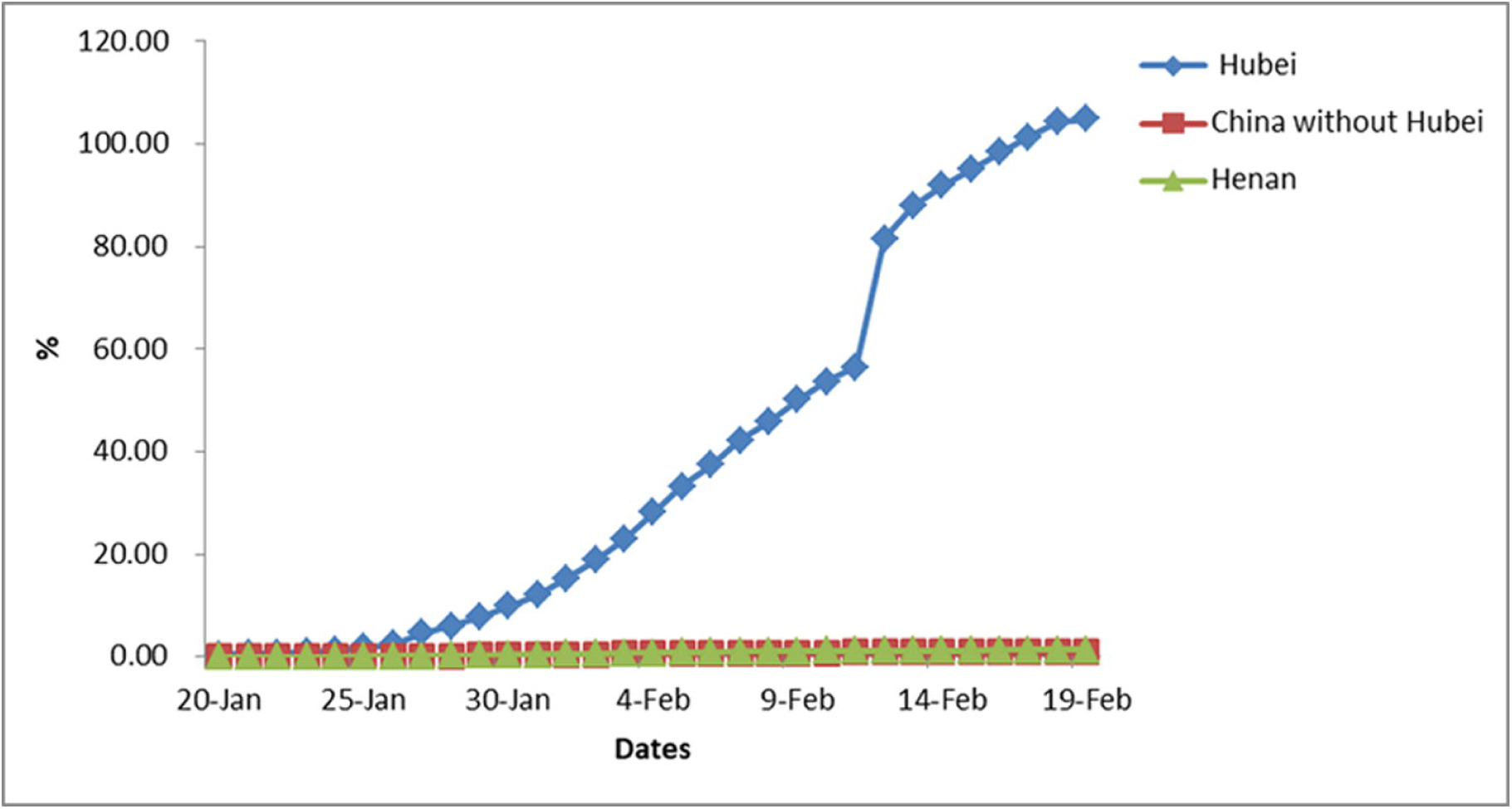
Cumulative incidence rate among various groups.

**Fig 2.**
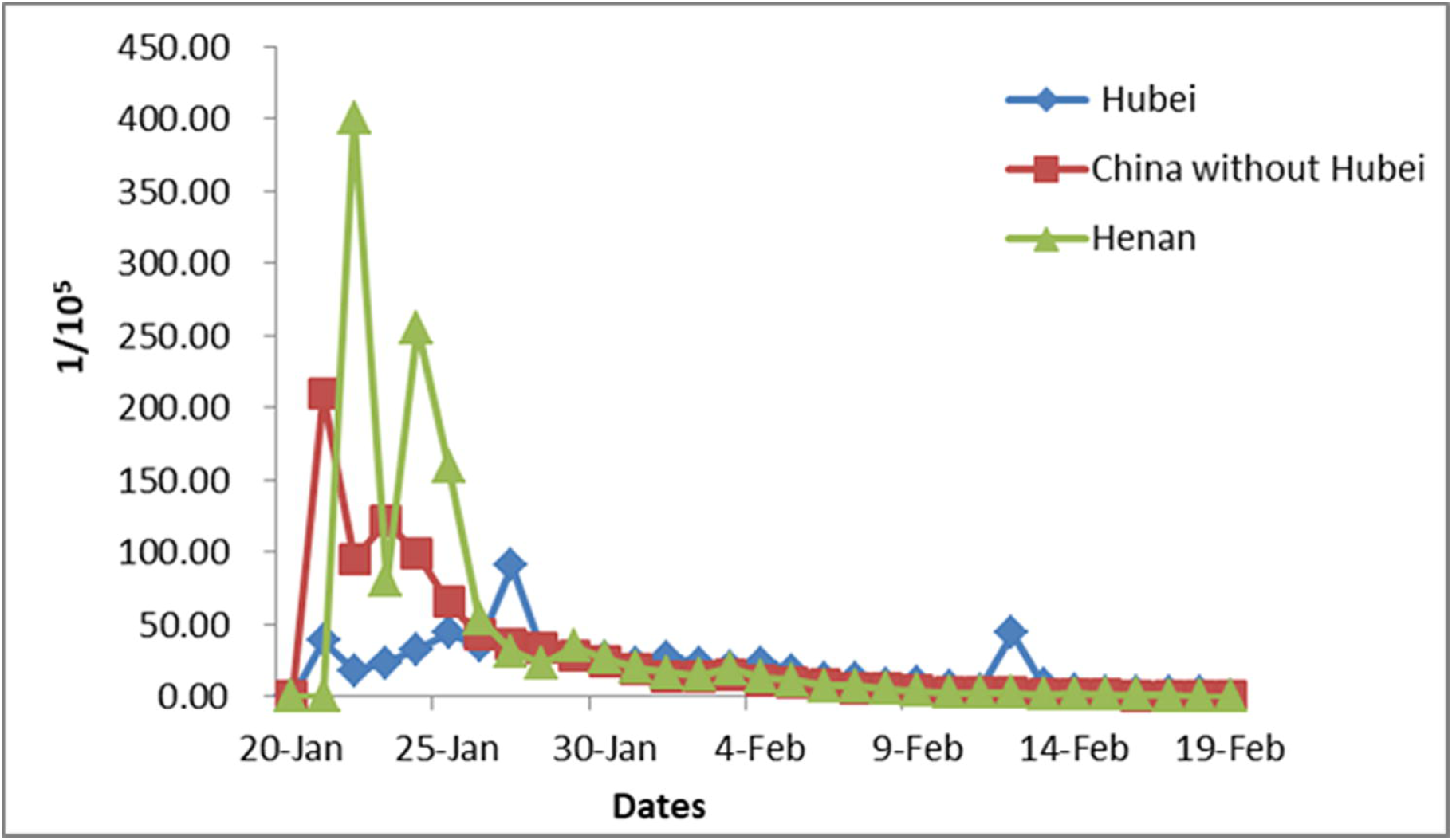
Chain growth rates among various groups.

According to known data, the best parabolic fitting was made for three groups (Figure 3, 5 & 7). With SIR model, a series of differential equations were solved, then fitting curves were made to approach to reality infinitely. In figure 3, the simulated curve in the Hubei Province fitted well with actual scatter plots. At beginning, the infected patients increases and reaches a peak with 51,673 cases around on February 18, then the curve decrease. After continually decreasing, the infected patients will maintain at lowest level in early May, suggesting the epidemic will eventually end (P<0.05). Figure 4 also shows that R_t_ change in Hubei Province with S type curve. It starts from 6.27 and decreases gradually to 1 around on February 18, and it is expected to approach 0 in the end of April and early May (P<0.05).

**Fig 3.**
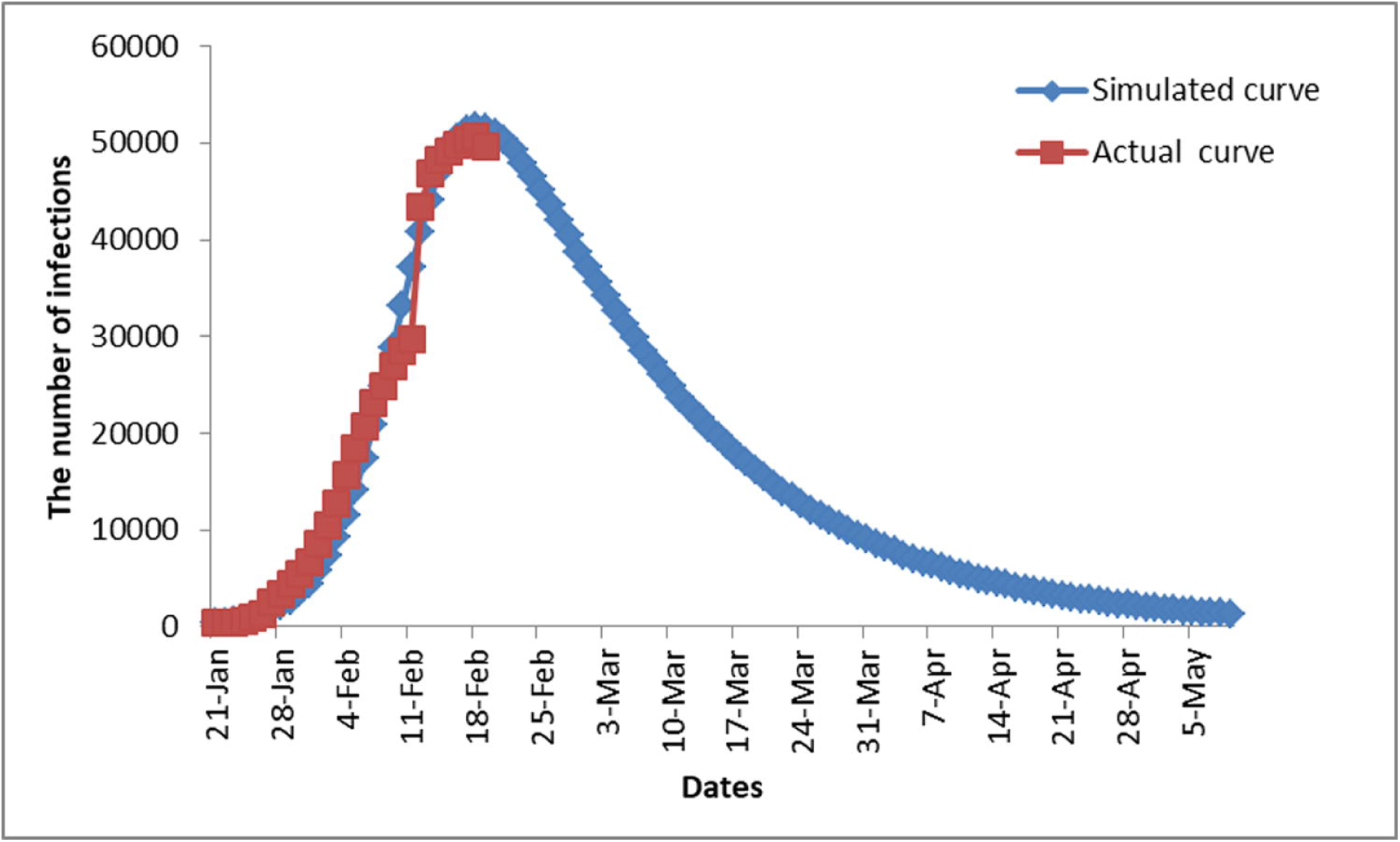
The actual curve and simulated curve in Hubei Province. The peak appears at 18/2, and the epidemic will end in May.

**Fig. 4.**
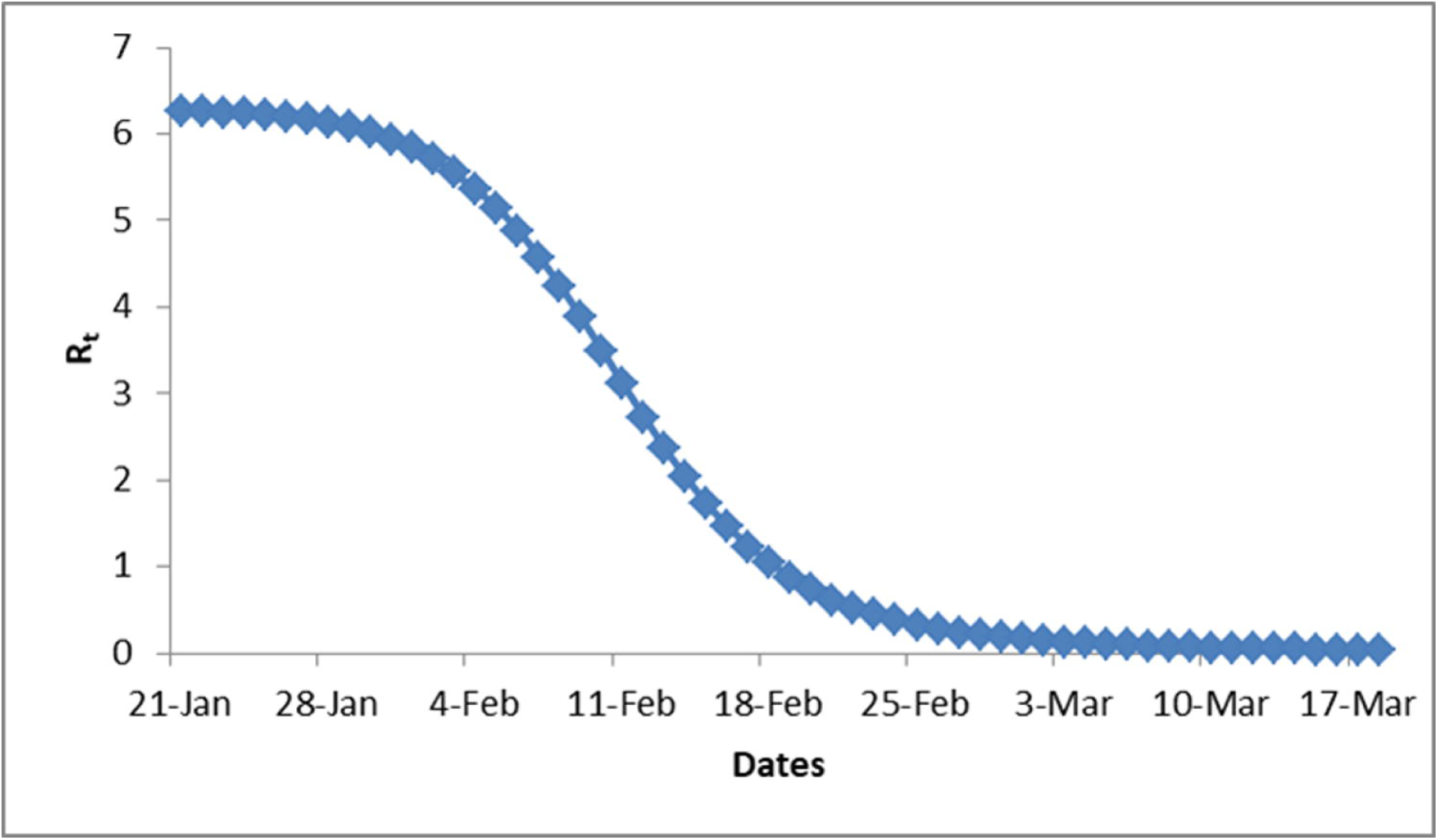
Dynamic R_t_ in Hubei Province with S type curve, and in early March, R_t_ closes to 0.

**Fig 5.**
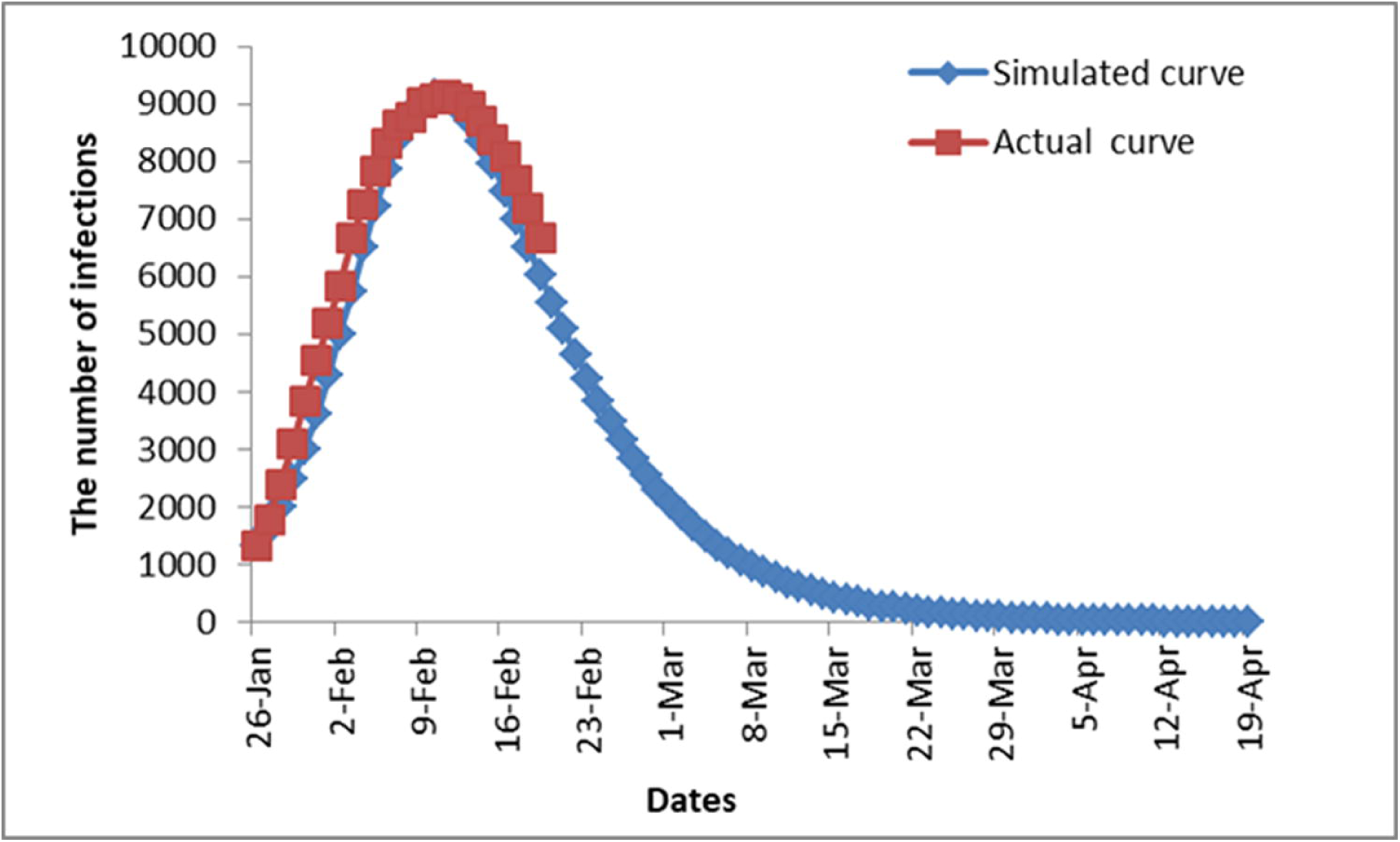
The actual curve and simulated curve in other areas of China. The peak appears at 10/2, and the epidemic will end in April.

**Fig 6.**
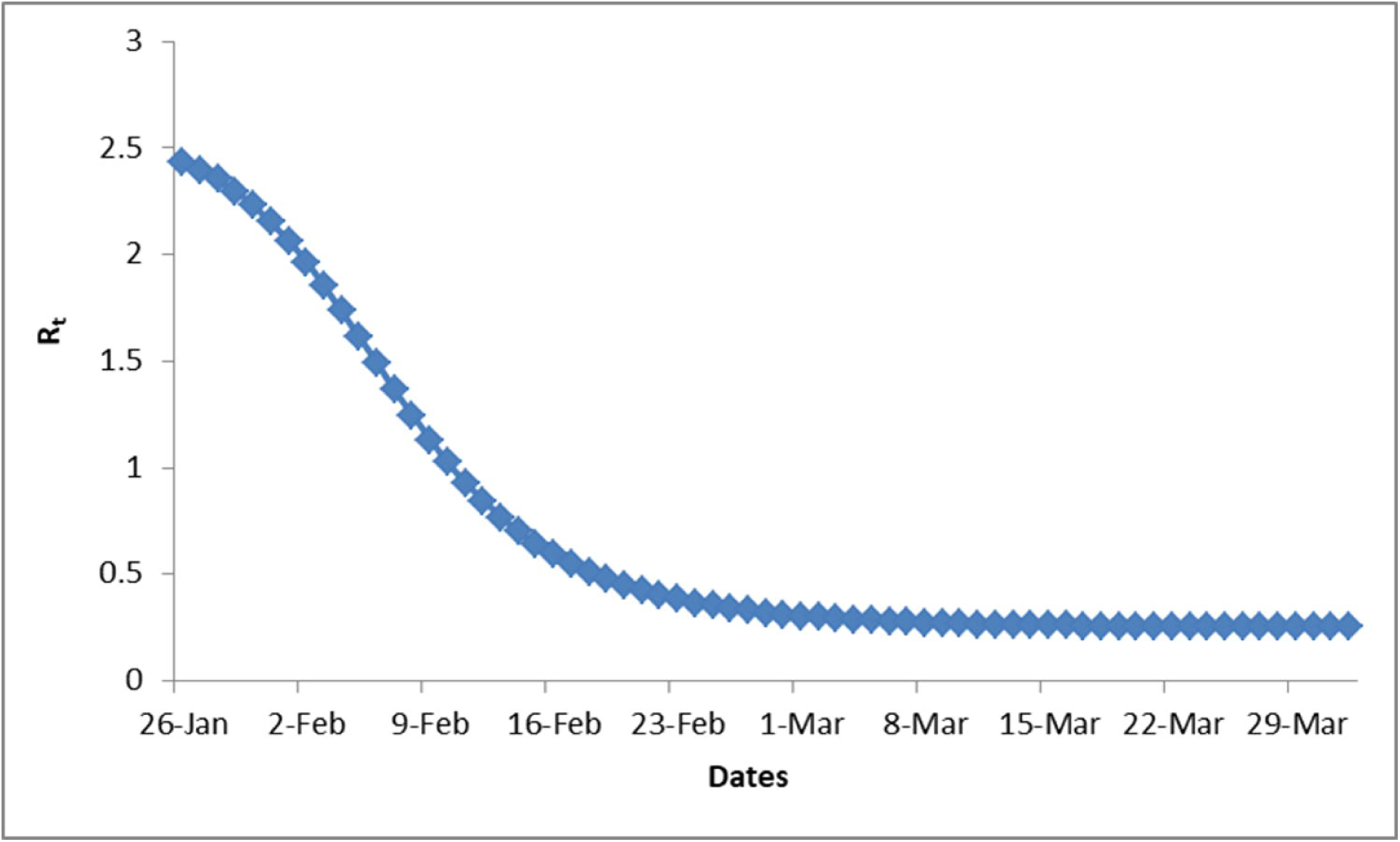
The dynamic R_t_ value in other areas of China with S type curve, and in the end of March, R_t_ approaches to 0.

**Fig 7.**
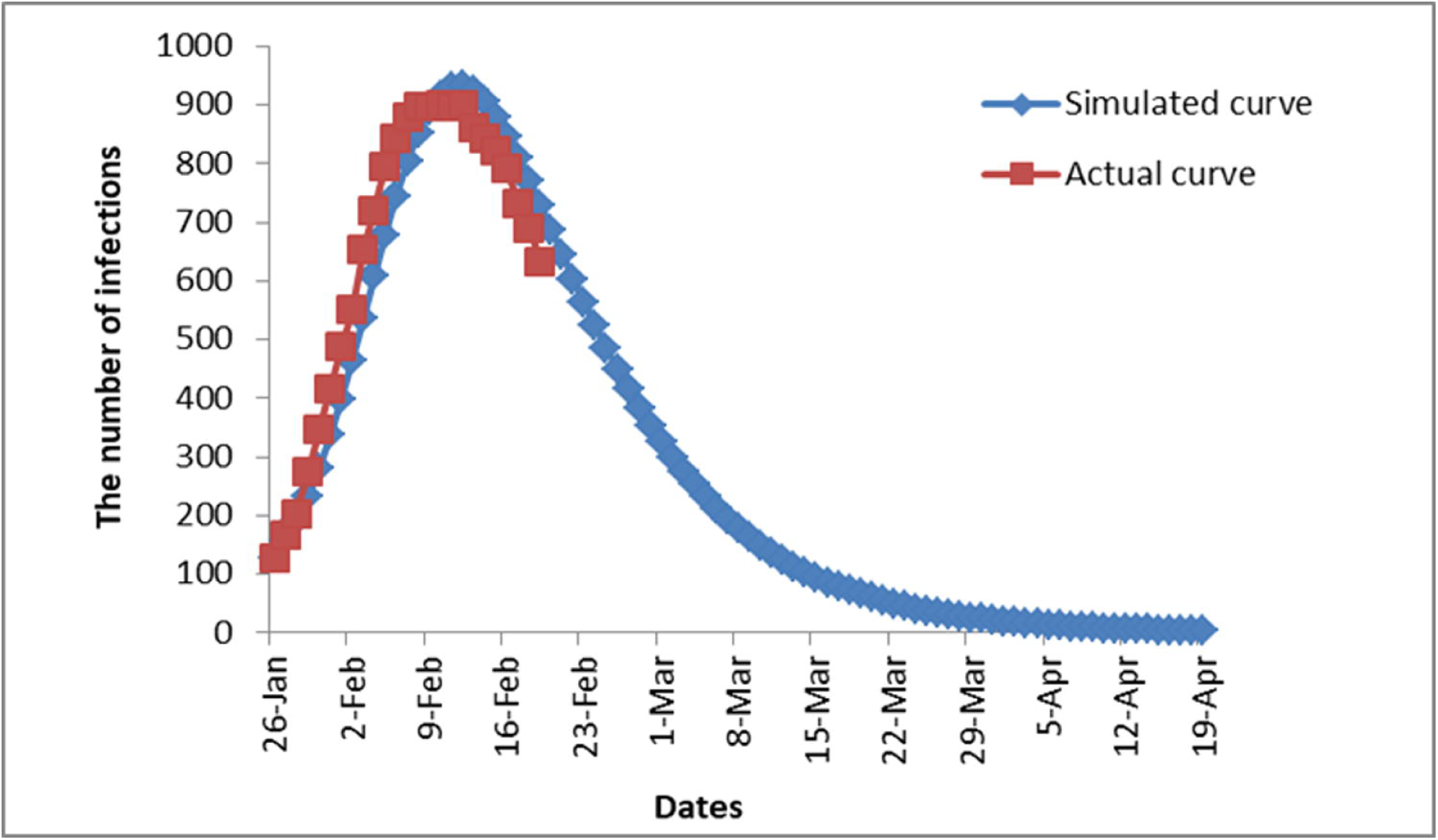
The actual curve and simulated curve in Henan province. The peak appears at 12/2, and the epidemic will end in March.

The parabolas were also fitted in the group of other areas of China and group of Henan province (Figure 5 & 7). The two groups were very similar, and the patient cases reach peak approximately on February 10, 2020. At peak, the infected patients are expected to 9145 cases and 931 cases respectively. After peak, the cases will gradually decrease, reaching the bottom in early April 2020. After that time, the epidemic will end. Figures 6 and 8 show that R_t_ values in nationwide without Hubei and Henan Province with S type. The values started at 2.44 and 2.9, and decreased to less than 1 after February 12th. They are expected to be close to 0 at the early April. Fig. 9 shows the infected patients among various groups.

**Fig 8.**
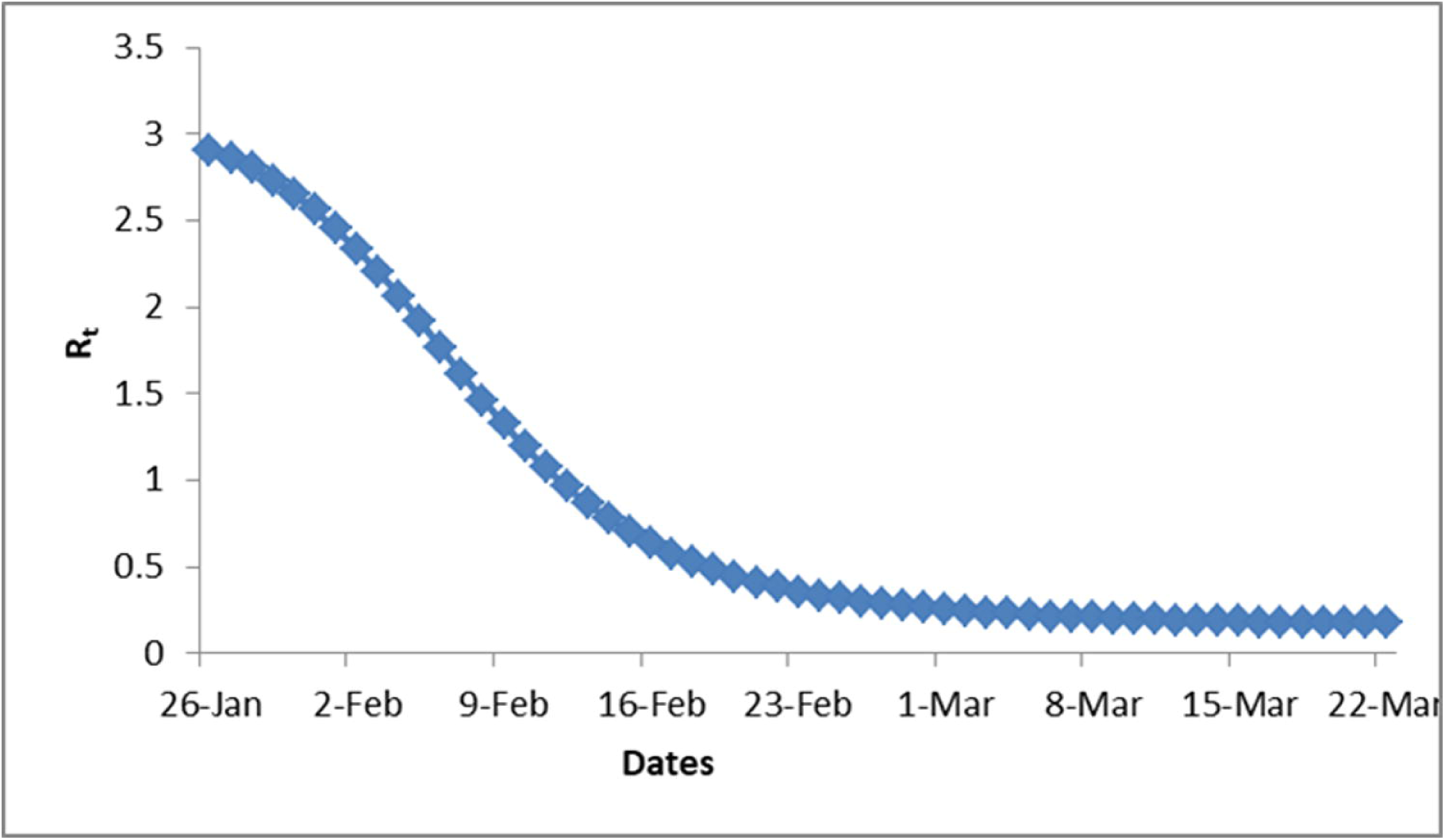
The dynamic R_t_ value in Henan Province with S type curve, and in the end of March, R_t_ approaches to 0.

**Fig 9.**
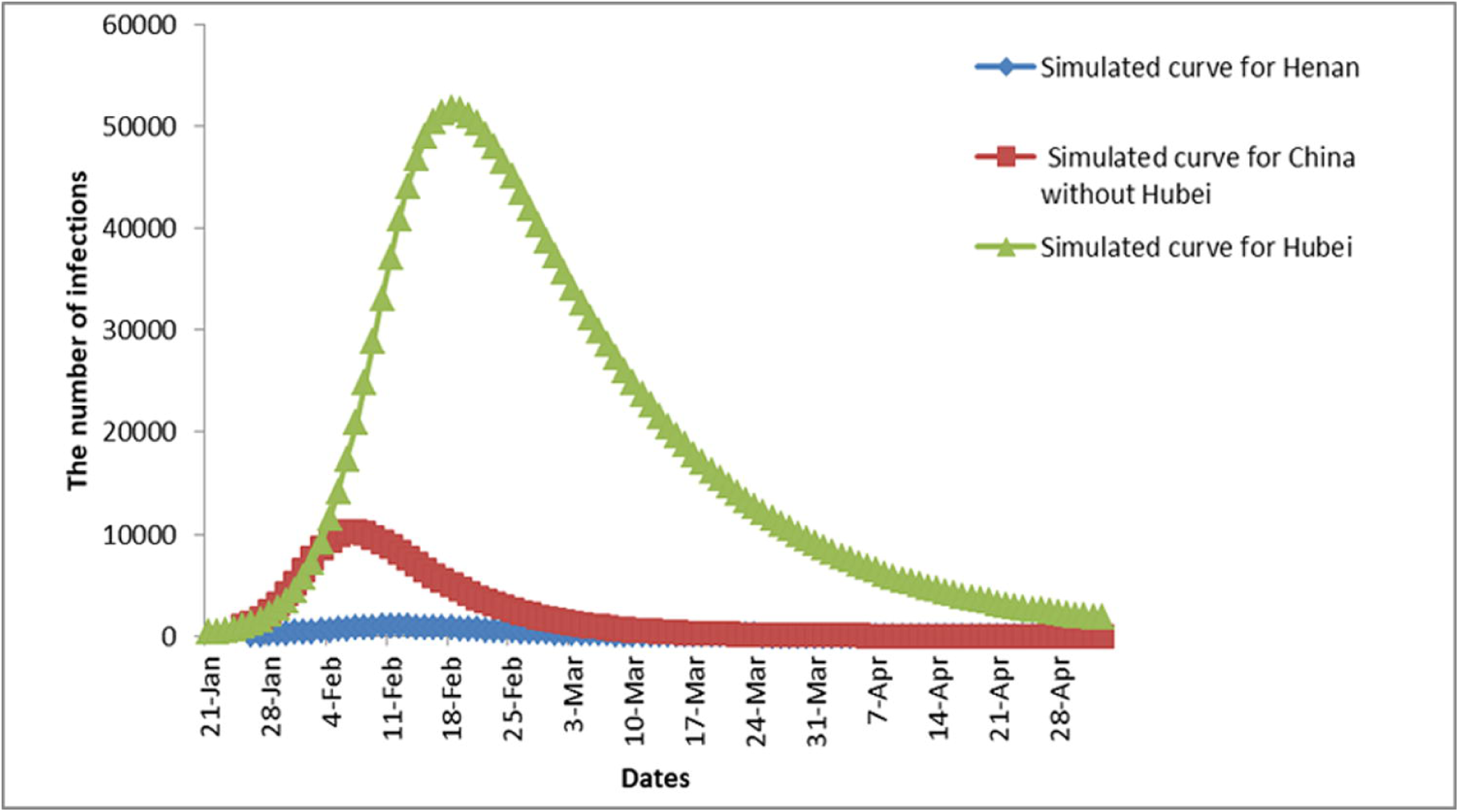
The infected patients among various groups. Simulated curves with blue, red and green colors represent Henan province, other areas of China and Hubei province respectively.

## Discussion

The epidemic development of COVID-19 shows a parabolic change. Since COVID-19 is highly transmissible and severely ill, scientists hope to understand its epidemiological development and forecast its future eagerly. Read et al. ever tried to calculate the disappear time of the epidemic.^12^ Since almost epidemics show their epidemiological development with classic parabolic distribution, therefore, SIR soft is designed according to the distribution. SIR is widely used to construct a mathematical model to understand the epidemic’s transmission and outcome of the diseases. Our results showed that COVID-19 followed parabolic distribution as well, but the infection, peak time and disappear time are different among various areas. Among the three groups, the patients reach their peaks approximately on February 10 (nationwide except Hubei) and February 18 (Hubei province) successively. Then the epidemic will disappear in early April (Nationwide without Hubei) and early May (Hubei province). Our estimation has some difference from Read et al. For instance, the peak value and cumulative cases seem to be less than theirs, and the ending time that we predict happens early. The differences may be caused by the data from different time and different source. Our data are relatively late and came from official publication. It should be acknowledged that our mathematical model is not perfect. Considering the sudden onset and some human intervention during the spread, our study only provides a rough description for the development of COVID-19 pneumonia.

R_0_ is valuable parameter to evaluate the epidemic transmission capacity. WHO has predicted that R_0_ of this novel coronavirus transmission ranged from 1.4 to 2.5.^13^ Using exponential growth model, R_0_ of SARS-CoV-2 in the early outbreak of Wuhan was estimated 2.24 (95% CI: 1.96-2.55) to 3.58 (95% CI: 2.89-4.39).^14^ With other methods, R_0_ was estimated to be 2-5,^15-17^ harmonizing with R_0_ of SARS-CoV (R_0_: 2-5),^18 19^ which is slightly lower than Middle East respiratory syndrome (R_0_: 2.0-6.7).^20^ In our study, R_0_ ranges from 2.44 to 6.27, higher than previous studies above, suggesting the strong transmission for the disease. After February 18, R_0_ goes to less than 1, suggesting the transmission becomes weak.

Hubei especially Wuhan is plagued by the epidemic. SARS-CoV-2 has 79.5% similar sequences.^21^ with SARS-CoV coronavirus and 51.8% similarity with MERS-Cov.^22^ but NCP’s sudden outbreak and rapid spread is much higher than SARS or MERS-CoV. For COVID-19’s outbreak, Hubei’s situation is the most serious in China. The infected patients is listed at No. 1 in the country, and by February 17 the cumulative cases are up to 59989. The epidemic also develops very quickly, and in early onset its rising curve is very steep. The cumulative incidence rate is much higher than other areas of China. According to the fitting curve, the peak will arrive on February 18, several days later than other areas in China. In the meantime, the disappear time of epidemic will delay until early May, one month later than other areas. The epidemic period with more than half year in Hubei also lasts much longer than other areas in China. On the other hand, in nationwide without Hubei and Henan province, the number of patients and incidence rate have been kept at a relatively low level. In terms of contagion, at beginning, R_0_ in Hubei is 6.27, much higher than 2.44 in other areas of China. In this way, we can conclude that Hubei is serious stricken regions of the epidemic. There are several reasons to explain these differences between Hubei and other areas. (1) Hubei is origin of the plague, and the earliest cases were found in Wuhan. It is these patients who cause the widespread. (2) In early epidemic stage, the disease’s jeopard and its infectious capacity have not been caused sufficient attention, therefore, the interventions are ineffective and inadequate. (3) Closed cities accelerate the disease transmission in Wuhan and Hubei, although the path of outward transmission is cut off. After January 23, Wuhan city and Hubei province started to close cities successively. In this way, about 60,000,000 people were limited in Hubei province. This makes Hubei become a closed infection system, which is bound to accelerate the spread of the disease. On the other hand, about 5 million people left Wuhan before the cities were closed, although the 5 million people became source of infection to other areas in China, the biggest source of infection is cut off. Fortunately, in Hubei, more interventions were accepted, for example calling on the citizens to stay at home.^23^ In this way, further transmission each other decreases, and the epidemic is slowed down. Due to reasons above, Hubei Province should be regarded as the plague hardest hit area and must acquire more medical supports. Luckily, more medical teams from all over the country have been sent to Wuhan city and Hubei province, and more and more effectively diagnostic kits and medicines can meet the Hubei’s needs gradually.

The epidemic forecast is important for NCP prevention. Our epidemiological model has guiding significance for COVID-19 prevention. At present, closed management of towns and communities is carried out in many areas throughout the country. Many traffic are blocked, and works, schools and markets are suspended,^24^ which has brought extremely inconvenience for citizens. Citizens look forward to a rapid improvement in this situation. Probably, our study is helpful to solve the problems above. The epidemiological model shows: from February 10 to 18, the confirmed patients will reach their peaks in various areas successively, and after that dates, the epidemic will be improved. R_t_ will also approach 0 after April in nationwide without Hubei and after May in Hubei province. It means that the epidemic will end without infection, suggesting the closed management can be lifted after May in Hubei and April in other areas of China, then citizens can return to normal work and life.

In summary, with classic SIR models, the fitting epidemiological curves have made to simulate the occurrence, development and end of novel coronavirus pneumonia in China, and R_0_ is calculated as well. Our study shows: (1) The epidemic development shows parabolic curves, and the incidence is expected to reach its peaks on February 10 in nationwide without Hubei and on February 18 in Hubei province successively. The epidemic will end after May in Hubei and after April in other area of China. (2) In early outbreak of epidemic, R_0_ was as high as 2.44-6.27, and with epidemic development, R_t_ will decrease and finally approach 0 after April in nationwide without Hubei and after May in Hubei province. (3) According to the data above, we suggest the closed management in city and community should be lifted after May in whole China. Without epidemic infection, citizens return to their normal work and life.

### Contributors

Shanshan Wu and Panpan Sun wrote the initial draft with the guide of Jinbo Deng, and Jinbo Deng edited to subsequent revisions. The first two authors contributed equally to this article. All authors provided critical feedback and approved the final draft of the manuscript. Jinbo Deng is the correspondent author who attests that all listed authors meet authorship criteria.

## Data Availability

Dissemination to participants and related patient and public communities: No study participants were involved in the preparation of this article, which permits others to distribute, adapt and build upon this article noncommercially.

## Ethical approval

This study was not invovled in ethical issues.

## Transparency

The process of data collection is transparent and rational.The lead authors affirm that the manuscript is an honest and scientific statement. and Any discrepancies from the study as planned have been explained.

